# Impact of reduction of susceptibility to SARS-CoV-2 on epidemic dynamics in four early-seeded metropolitan regions

**DOI:** 10.1101/2020.07.28.20163154

**Authors:** T. J. Barrett, K. C. Patterson, T. M. James, P. Krüger

## Abstract

**As we enter a chronic phase of the SARS-CoV-2 pandemic, with uncontrolled infection rates in many places, relative regional susceptibilities are a critical unknown for policy planning. Tests for SARS-CoV-2 infection or antibodies are indicative but unreliable measures of exposure. Here instead, for four highly-affected countries, we determine population susceptibilities by directly comparing country-wide observed epidemic dynamics data with that with their main metropolitan regions. We find significant susceptibility reductions in the metropolitan regions as a result of earlier seeding, with a relatively longer phase of exponential growth before the introduction of public health interventions. During the post-growth phase, the lower susceptibility of these regions contributed to the decline in cases, independent of intervention effects. Forward projections indicate that non-metropolitan regions will be more affected during recurrent epidemic waves compared with the initially heavier-hit metropolitan regions. Our findings have consequences for disease forecasts and resource utilisation.**

## Introduction

Coronaviruses have long been an endemic source of human and animal infections. SARS-CoV-2 appeared in late 2019 [1–3], causing the potentially fatal illness COVID-19. Due largely to human mobility patterns and infection characteristics, SARSCoV-2 became a pandemic [4], which continues to wreak public health, social, and economic havoc across the globe. While a variety of strategies have been implemented to attempt to control the spread of disease [5, 6], the death toll continues to rise and infection rates are uncontrolled in many parts of the world.

Knowing the extent of population exposure to SARS-CoV-2 has important implications for public policy measures, including ongoing efforts to contain infection [7]. Yet, while SARS-CoV-2 has an extensive global reach, the portion of populations currently and previously infected remains unclear. Viral assays only capture recent infections, and even then can be unreliable. Serological antibody testing to ascertain levels of prior exposure is burdensome, with unreliable testing validity and the potential to lead to systematic under-counting [8, 9]. Tracking epidemic data across populations and evaluating the time evolution of key markers such as death rates provides a powerful alternative devoid of these shortcomings.

For this analysis, we evaluate the growth and declines of case numbers during the well-defined epidemic phases. Generally, when a fully susceptible population is exposed to a new and contagious pathogen, an epidemic starts with an initial seeding phase when a small number of individuals are infected. Unconnected local outbreaks with unpredictable spread can occur due to variability in individual behaviour, the effects of potential ‘super-spreading’ events, and the non-uniform success of containment measures. This is followed by a phase of free exponential growth, during which ‘track and trace’ based containment is no longer feasible. Exponential spread of infections *I*(*t*)=e^(^*^β^*^−γ)^*^t^* over time t happens at a rate 1/*τ* = *β* − γ> 0, when the average rate of new secondary infections caused by each primary infectious individual (*β*) exceeds the rate of recovery (γ).^1^ By reported daily death counts [10], the disease dynamics of COVID-19 have fulfilled the condition of exponential growth and decline in nearly all parts of the world. For four countries highly affected by COVID-19 and with reliable reporting indices, we utilise this epidemic data to estimate the fraction of sub-populations which remain susceptible to SARS-CoV-2, which is a key parameter governing the course of the epidemic.

## Results

While onset of the exponential phase varies due to different seeding times, we find that the time constant of exponential growth is very similar over a wide geographical range. Fig.1a illustrates this using data from four highly-affected countries: Spain, Italy, the United Kingdom, and the United States [11].

**Figure 1:**
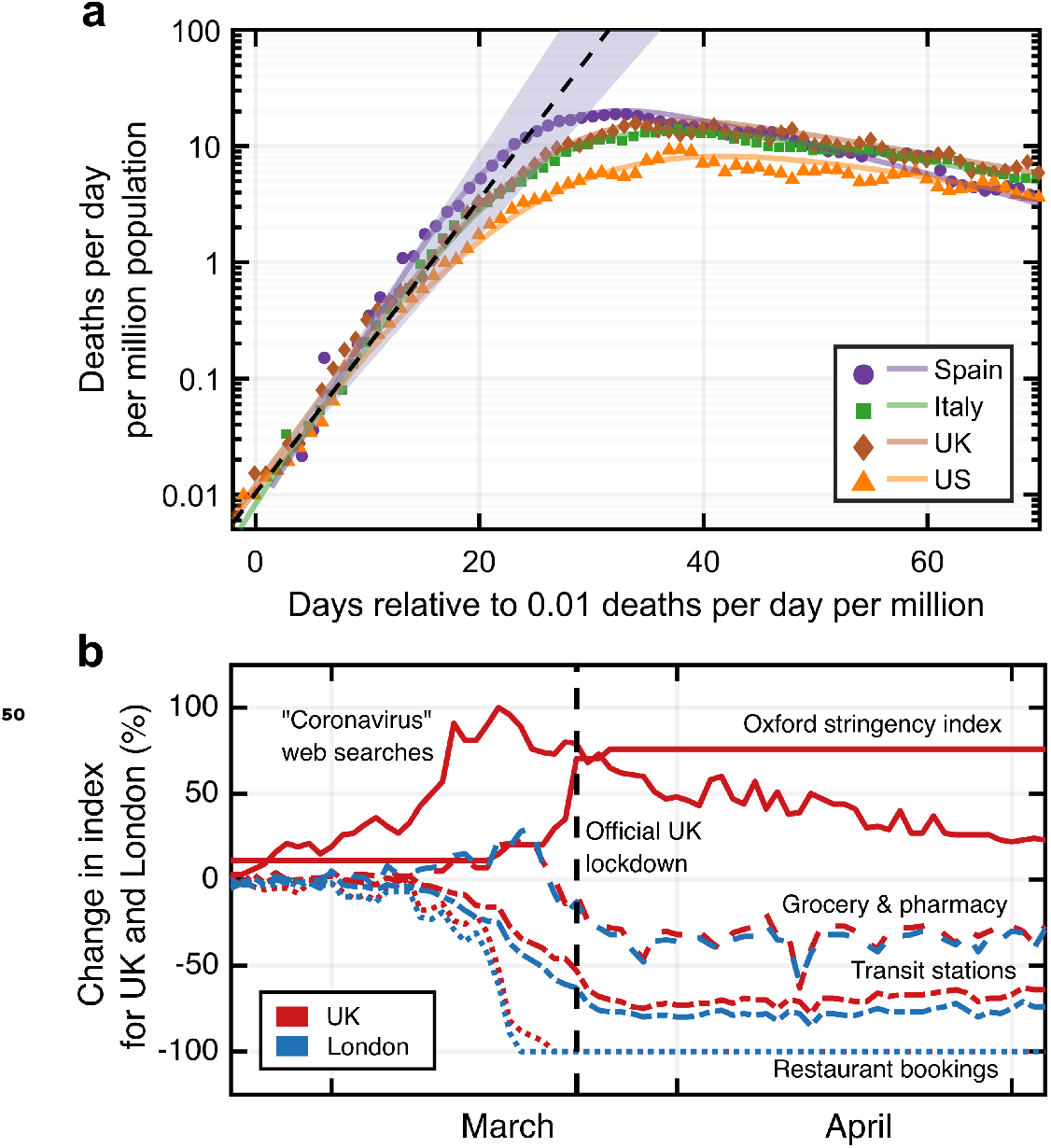
a) Reported number of daily deaths (smoothed by a moving average over 7 days) in four highly affected countries. The dashed line (shaded area) represents the mean (range of) early exponential growth(s) with a time constant *τ* =3.4(2) days^2^. The lines are spline interpolations to guide the eye. b) Comparison of different measures that potentially affect infection rates, shown for the example of the United Kingdom.

The exponential growth is sustained as long as *S*, defined as ratio of susceptible individuals to the whole population, is close to 100%, and *β* and γ are kept constant. Slowing spread and ultimately reversal to an exponential decline can be driven by an increase of γ, through interventions such as anti-viral therapies that reduce infectiousness time. Such therapies are not currently available for COVID-19. Consequently, reductions of new COVID-19 infections can only be attributed to reductions in either *β* or *S*. Public health interventions aim to reduce *β* through hygiene, social distancing, and mobility restriction measures designed to reduce contact frequencies [5, 6, 12]. In the following analysis we argue changes in COVID-19 disease dynamics are due to reductions of *β* and *S*, with different relative importance across geographic areas. In particular, a significant reduction of *S* is observed in the main metropolitan regions of the four countries investigated here.

### Observed impact of susceptibility reduction on infection dynamics

Variability in the nature and timing of public health interventions and resulting population behaviours are eventually reflected in variations across countries in the slowdown and exponential decline of the daily death counts. Fig. 1b illustrates, for the specific example of the United Kingdom and London, the timelines of public awareness (measured through frequencies of relevant internet search queries [13]), government restriction stringency (measured as aggregate ‘Oxford’ stringency index [14]), and visits to public meeting places (restaurants [15], grocery stores, pharmacies, and public transit stations [16]). Qualitatively, all measures correlate with one another, with both voluntary behavioural changes and the stringency of public health interventions resulting in reduced frequency of visits to locations of high infection risk.

While it is difficult to quantitatively associate a time-dependent reduction of *β*(t) with these data, the observed location data indicate that the public response between areas within one country is similar in timing and magnitude of deviation from baseline. Due to the seeding of the epidemic in each country’s main metropolitan region (Italy - Milan/Lombardy^3^; Spain Madrid; England - London^4^; USA - New York City) occurring earlier than in the rest of the country, the time between epidemic onset and reduction of *β* (driven by public behaviour change) differs in an otherwise comparable situation. We therefore consider pairwise comparisons of main metropolitan regions and rest of countries a suitable tool to investigate the relative impact of reductions in *S*.

Fig. 2 (upper panels) shows reported daily death counts [11, 17] for the four investigated countries, broken down into the data from the main metropolitan region and the rest of the country. Each of the corresponding eight data sets shows an initial exponential growth phase, followed by a transition to an exponential decline. Seeding of the epidemic consistently happened earlier in the metropolitan regions than elsewhere in each country, leading to higher population-normalised daily death counts in these areas. Yet, the time constant of the exponential growth is equal within errors for each metropolitan-country pair (except in the United States due to its heterogeneity), suggesting that the contact rate among a fully susceptible population is not critically dependent on factors related to population density [18, 19]. In contrast, the subsequent decline is consistently but to a varying extent faster in the metropolitan regions than in the rest of the country. Together with the notably earlier departure from the exponential growth phase and the observation that public policies within countries were similar, this indicates that the reduction of *S* played a more important role in the metropolitan regions relative to the rest of the country in all four cases.

**Figure 2:**
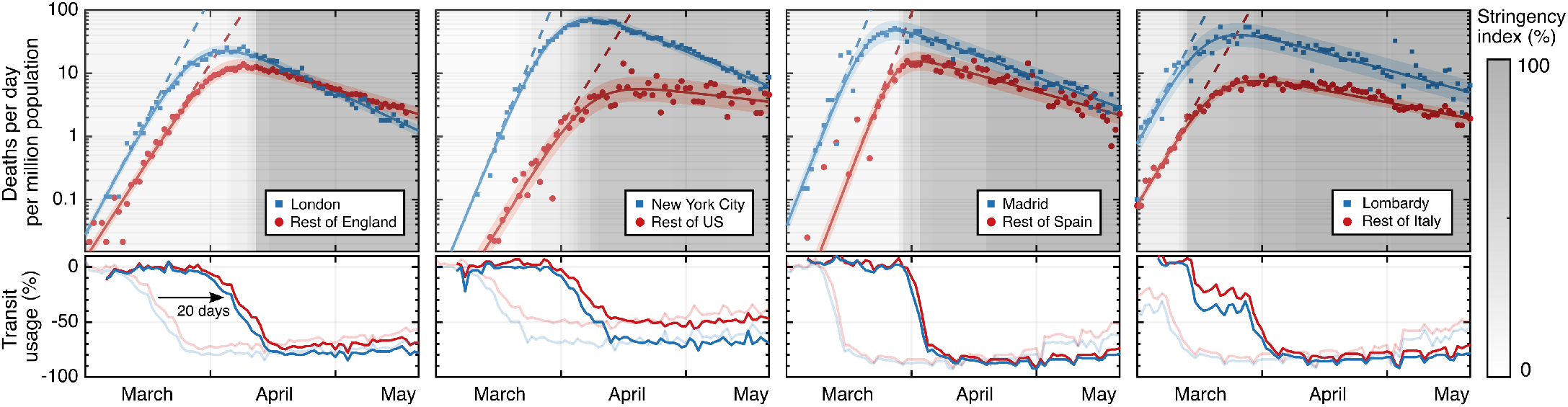
Population-normalised daily death counts (logarithmic scale) of most populous metropolitan region (blue) and surrounding country (red), together with fits and confidence intervals shown (see Methods for details). The stringency index, time shifted to account for an average delay between exposure and death, is grey-scaled in the background. Public transit station use (bottom panels) is shown for comparison, with faded lines for real-time data and bold lines for time-shifted.

Quantitatively, the time constants *τ_m_* and *τ_c_* of the exponential decay phase provide a relative measure of the reduction of susceptibility in each metropolitan region (*S^m^*) compared with the remainder of the country (*S^c^*). The observation of an extended period of purely exponential decline, together with a stable interval of public health interventions, shown in Fig. 1b and Fig. 2 (lower panels), (even when taking into account a delayed impact of ≈ 20 days of their effect on the daily death counts [20, 21]), implies that both *β*(*t*) and *S*(*t*) do not vary much during this phase. In this case, the decay time constants can be used to yield the ratio of fractions of the population that remain susceptible to the virus, provided *β* = *β*_m_ = *β*_c_ remains equal across each country [22]. From the fits presented in Fig. 2, we find for the ratios *S^m^*/*S^c^* values of 0.86 ± 0.05 (Madrid/Spain), 0.89 ± 0.04 (Lombardy/Italy), 0.70 ± 0.06 (London/England), and 0.59 ± 0.05 (New York/United States). These ratios represent upper bounds for *S^m^*, since *S^c^* is by definition ≤ 1. Note that if instead *β^m^* ≠ *β^c^*, then the ratio of susceptibilities will be corrected by a factor *β^c^*/*β^m^* (see Methods).

We directly compare the relative timing of the epidemic evolution with public policy and behaviour changes (Fig. 2). The results corroborate the finding that a reduction of *S* had already begun to reduce new infection rates in the metropolitan regions when public behaviour changes started to have a significant impact. The difference in the effect of *S* for all eight geographic areas is shown in Fig. 3, where the peak number of daily deaths is plotted against the time delay between when visits to public meeting places are reduced and when death rates begin to slow (see Methods). The country-level data cluster around a common *delay time* that is similar to the reported average time between viral exposure and death [20, 21]. This suggests that the main driver for the initial reduction in death (infection) rates is changes in public behaviour. This is not the case for the metropolitan region data, which in turn cluster around a common *peak daily death count*. This clustering suggests that the larger early spread of the disease was sufficient to reduce infection and death rates through reduction of *S* in addition to the effects based on behavioural changes.

**Figure 3:**
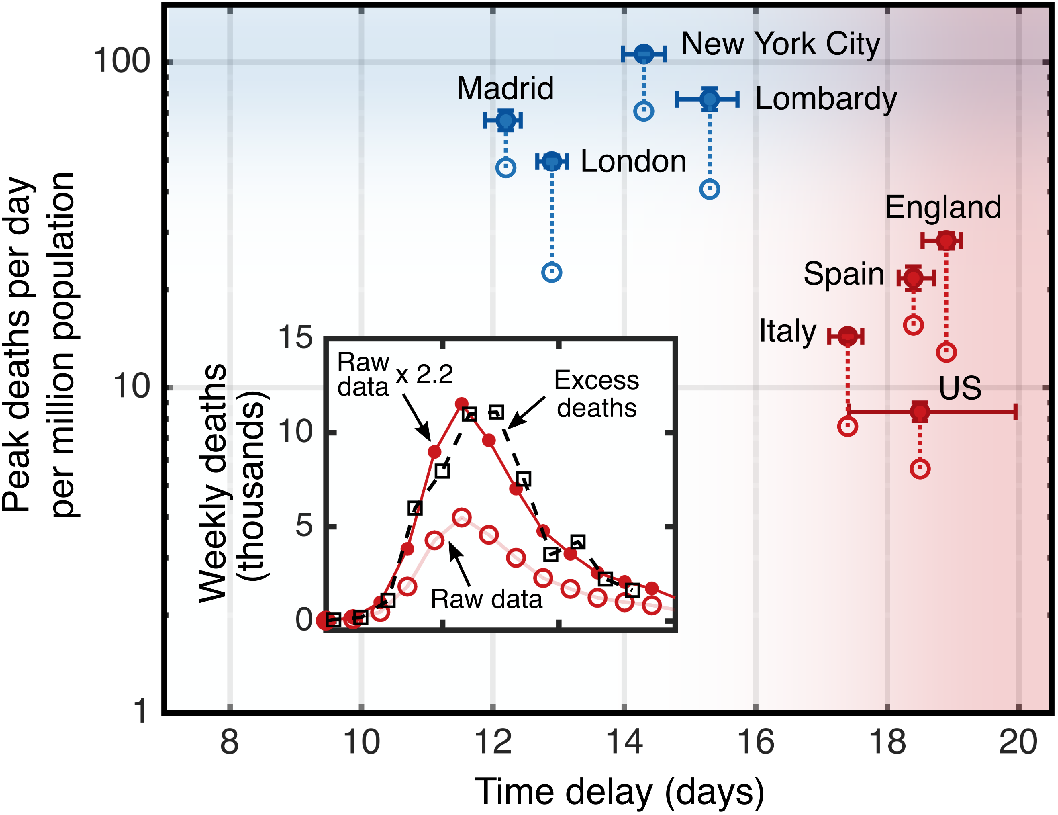
The peak daily death counts plotted against the days between public behaviour change and the end of exponential growth leading to a peak rate. A correction factor from excess death data was applied to the reported rates (open circles) to derive the expected rates (solid circles), which are linked. The 4 countries exclude the corresponding metropolitan regions. The inset illustrates, for the example of England, a death count calibration by comparing reported COVID-19 deaths to the all-cause excess death data.

### Modelling and forecasting

To further investigate the dynamics of the epidemic in each area, we employ the well-established compartmental Susceptible-Infected-Recovered (SIR) model [23, 24], and solve the differential equations simultaneously for each pair of country and metropolitan regions, with coupling between the two (see Methods for details). The solutions for each area are used to estimate *S* and the time dependence of it, as well as to explore the potential future evolution of the infection dynamics. We incorporate public behaviour changes into the system by adjusting the trajectory of *β*(*t*) equally for each metropolitan region and country pair, informed by Fig. 2 (lower panels). To evaluate the effect of initial public health measures on S, we only include the death counts until mid-May 2020, to avoid any effects of the relaxation of restrictions on the data.

In contrast to the model-independent slope comparison from the previous section, solving the model equations also provides absolute values for the susceptibility and infection fatality rate *µ* in each area, which are shown together in Table 1. Using both approaches, we consistently find a comparatively weaker reduction in *S* outside the early-seeded metropolitan regions.

**Table 1:**
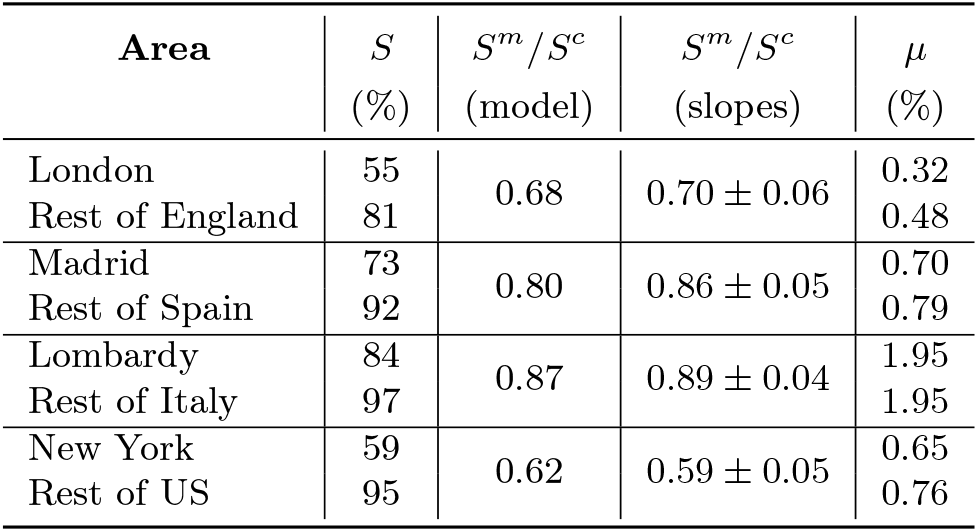
Results of solving the SIR model with a time-dependent contact rate in each area. Ratios of susceptibilities are shown, along with the model-independent approach of analysing slopes for comparison. The model also provides absolute values of susceptibility *S* and infection fatality rate *µ*.

Figure 4 shows the result of the model for the specific example of England and London, demonstrating that public policy interventions helped cap the death counts in both cases, but that a reduction of *S* is also a significant driver, particularly in London. Without relaxation of initial interventions, containable infection levels (set at ≾ 10 new daily cases per million population^5^) would be projected to occur in mid-July and mid-August in London and the rest of England, respectively (solid lines **c** in Fig. 4). In reality, a relaxation of restrictions (early-June 2020) and easing of travel restrictions have been initiated. As a result, *β* increased, and slowed down the decline in death rates. In a forward projection based on a continuation of these new conditions with no further changes to *β*, containable infection levels in London and the rest of England could be reached in mid-August and the end of 2020, respectively (dash-dotted lines **d_2_**). As further relaxations are underway, these dates may be further delayed if no compensating changes in public behaviour are introduced to maintain the early-July value of *β* =0.135 d^−1^. We note that this value is already greater than the observed value of γ (see Methods), and thus would have been enough to drive another period of exponential growth in the absence of any reduced susceptibility effect.

**Figure 4:**
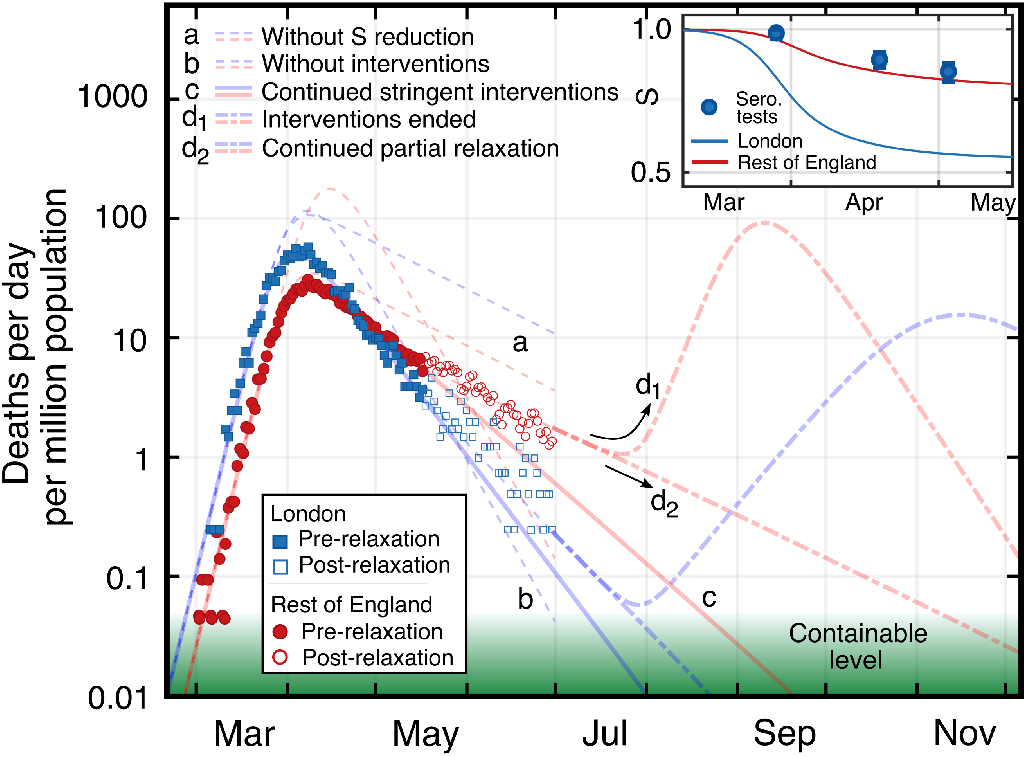
SIR model and forecast for London (blue) and the rest of England (red). Adjusted daily death counts are shown pre and post relaxation of interventions. The hypothetical scenarios of no reduction in susceptibility *S* (a) and of no interventions (b) are displayed. Extrapolations for scenarios of continued stringent interventions (c), of ending interventions (d_1_), and of a partial relaxation of stringency consistent with post-relaxation observations (d_2_) are shown. Inset shows the reduction of *S* for London and the rest of England, together with fractions of positive serology testing for London.

We also model the hypothetical scenario of a full return to pre-intervention conditions, constituting the second wave (dash-dotted lines **d_1_**). In this projection, infection rates begin to rise but the second peak for the metropolitan regions is reduced with respect to the first peak, while the second peak for countries is of higher magnitude than the first one, which is because the height of the peak scales with *S*.

In Fig. 4 (inset) the resulting evolution of *S*(*t*) for London and the rest of England is shown, together with the results of serology antibody tests [25], which have recently been suspected to underestimate the actual (yet potentially temporary) immunity levels due to the lack of sensitivity of the tests to T-cell based immunity [26]. In both methods, the metropolitan region exhibits a lower absolute value of susceptibility when compared with the rest of the country.

## Discussion

We have found that the dynamics of the SARS-CoV-2 pandemic have been influenced both by policy interventions and by reduced population susceptibility, with a relatively stronger contribution from susceptibility changes in early-seeded metropolitan regions. Our data provide an upward revision to the reported prevalence estimates derived from laboratory testing. In addition, while infection and thus death rates are a function of *S* and *β*, the effect of *S* reduction is more durable, and *β* will rise when restrictions are relaxed unless protective measures prove to be fully compensatory. If they are not, a second peak may occur. The reduction in *S* would not necessarily avoid this, but would mitigate peak height. Forward projections based on the observed data also indicate that later seeded areas are relatively more vulnerable in a second wave, which has implications for the distribution of healthcare resources.

Our model assumes a homogeneously mixing population. Yet, real-life infection spreading is heterogeneous, with some subregions of cities or neighbourhoods more heavily affected than others, as well as demographic or geographical groups mixing more strongly internally than between each other. As a result of these heterogeneities, susceptibility-mediated effects on epidemic dynamics can occur at lower exposure levels than would be required for a homogeneously mixed and exposed population [27]. As a consequence, our absolute estimates of remaining susceptibilities in each region may need to be interpreted as *effective* values that cannot be used to directly infer cumulative fractions of the populations having been infected with the virus. For example, the figures for *S* reported in Table 1 cannot be read as 19% (45%) of the population in London (rest of England) have been exposed to SARS-CoV-2. Instead the observed epidemic evolution in the heterogeneous population follows a pattern that would be expected for a hypothetical homogeneous population in which the exposure rates had reached these levels. The *ratios* of effective exposures between the areas are likely to represent the true exposure ratios. For example, we find that the fraction of the population that has been exposed to SARS-CoV-2 is 2.4 times greater in London than in the rest of England. The recent REACT study of antibody prevalence finds a very similar ratio of 2.2 between London and (total) England exposure (including London). While our absolute effective exposure levels are higher than the actual levels, the REACT study [28] could underestimate current immunity levels by a factor of two or higher, as T-cell based immunity can exist with negative laboratory antibody response [8]. The use of sub-standard finger-prick blood samples, as well as testing bias introduced through inhomogeneous test responsiveness in the selected test population, are further limitations of the REACT study.

Our modelling also assumes lasting immunity following acute infection, in keeping with normal immunological responses to viral infections [29]. Yet, for the purposes of our projections, immunity need only be preserved for as long as infection rates are uncontrollable. The level used for ‘controllable disease’ was chosen in accordance with data from countries achieving containment, and assumes a goal of elimination utilising, among other measures, tracking and tracing. It is possible that technological improvements will allow a successful track-and-trace strategy at higher infection levels. Forward-projecting continuations of modest relaxations of restrictions appears to make containment achievable, but observed slow down in the decline of death rates indicates that this will be reached later than would have occurred under more stringent restrictions.

## Data Availability

All data are freely available in publicly available repositories. Analysis code available upon request.

## Acknowledgments

We would like to thank R. Thorne for insightful comments and helpful discussion on the manuscript.

## Methods

### Extraction of epidemic parameter values

The key parameters of an epidemic can be extracted from epidemiological data (infection, hospitalisation or death rates), as long as populations are sufficiently large for demographic variations to average out, reported data are reliable, and interventions are relatively uniformly implemented. The similar observed exponential growth illustrates that this condition is fulfilled for the four countries of our study (the United States is an exception in some respects due to the heterogeneity of interventions and population behaviour). A key quantity is the time evolution of number of infected individuals. Due to insufficient quantity and quality of testing however, reported daily deaths *D*(*t*) are likely a more reliable measure of COVID-19 pandemic dynamics than the number of confirmed cases. The infection fatality rate (IFR) *µ*, assumed to be a constant with time, is the probability that an infected individual dies from the disease and therefore validates *D*(*t*) as a proportional measure of infections, and our main findings do not depend on knowing the exact value of *µ*.

For our investigated areas, *D*(*t*) is obtained from [11, 17]. Data for metropolitan regions is directly extracted from these sources, whereas data for the remainder of the country is derived as the national minus metropolitan data. Normalisation to the populations is based on the figures reported in Ref. [30].

The mean time constant *τ_i_* characterising the initial exponential infection growth, when the population is assumed to be fully susceptible, across the four countries is measured to be *τ_i_* = (3.4 ± 0.2) days. Using the clinically determined serial time *T_s_* = (4.0 ± 0.4) days [31, 32], i.e. the average time between primary and secondary infections, we obtain the basic reproduction number *R*_0_ = *β*/γ = exp(*T_s_*/*τ_i_*)=3.2 ± 0.3, which is consistent with the literature [6, 33, 34]. It follows that the contact rate is *β* = (0.43 ± 0.02) d^−1^ in an unconstrained population, and the recovery rate is γ = (0.130 ± 0.013) d^−1^, corresponding to a mean infectious period of (7.7 ± 0.7) days, again consistent with the literature [35, 36].

Turning now to the exponential decay phase, if the fraction of population that is susceptible *S* is instead different from 1, and has some other constant value, then the contact rate is simply modified accordingly and the final characteristic exponential decay time is given by

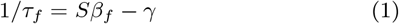

provided *β_f_* is also constant (for example during a period with little or no change in public behaviour). This allows us to examine the ratio of susceptible populations between a metropolitan region (m) and the rest of its corresponding country (c) through the relation

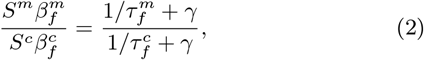

by measuring the time constants of the exponential declines. If *β_f_* is further assumed to take the same absolute value in the two areas, this equation simplifies to

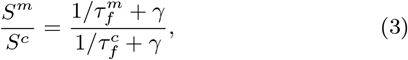

providing a direct way to estimate the ratio of relative susceptibilities in each area.

### Relating public health interventions (PHIs) to epidemic data

Variations in the stringency of country-wide public health interventions among countries have been extensively analysed by the Oxford group [14]. These data are not available at a regional (sub-country) level, and are not a direct measure of actual public behavior. We use cellular device location data [16] to overcome both these limitations, and to evaluate the effect of PHIs. Such data are available for several high infection risk location categories, including *retail and recreation, groceries and pharmacies, parks, transit stations, workplaces*, and *residential*. We plot these location data to show the relative change compared to the baseline value from the first five weeks of 2020 for each of the eight areas studied. Due to similarities of the relative timings and changes in magnitude of these measures, in principle any of them could be used as proxy for *β*(*t*). Based on the comparably low noise level as well as the consistent and clear functional shape of the location data at public transit stations, we chose these data for our analyses. Transit data, shown in Fig. 2 (lower panels), includes use of subway stations, taxi stands, sea ports, and other travel-related locations.

Empirically we find that the transit station location data are well described by a sigmoid function

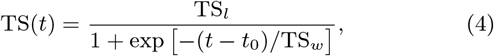

with a zero reference level (by definition the pre-pandemic normal level for early times *t* ≪ *t*_0_). The remaining fitting parameters characterise the magnitude of the response (TS*_l_*), the time (*t*_0_) when the public behaviour change happened and time duration (TS*_w_*) over which the change occurred. As a comparable measure of the time when a significant reduction of *β* is expected to occur, we choose the time *t*_50%_ when TS(*t*) is reduced by 50% from normal use as determined by the fit for each area, with the error determined as the one standard deviation functional prediction interval.

As the time evolution of *D*(*t*) follows exponential trajectories (growth or decline) during periods of (near) constant parameters *β*, γ and *S*, we display *D*(*t*) on a logarithmic scale and apply fit functions to *D*_log_(*t*) = log[*D*(*t*)] to avoid heteroscedastic bias. For fitting, we restrict data to a time window starting with exponential growth and ending just before the introduction of the first relaxation in PHIs where an uptick in TS(*t*) is observed (mid-May 2020). As a model function for fitting we use

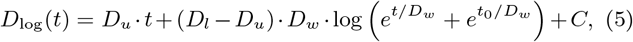

which represents an interpolation between two linear functions, and corresponds to an integrated sigmoid. The parameters *D_u_* and *D_l_* represent the values of the upper and lower linear slopes, respectively, and the temporal width of the transition between them is characterised by *D_w_*. We the fit this model to *D*_log_(*t*) in two steps. First, we individually fit the data at early and late times to a linear function using ordinary least-squares regression to obtain *D_u_* and *D_l_*. These values are then used as fixed parameters of the full function in a second fitting stage. We use the full fits to extract both the peak value of the fit to *D*(*t*) and the time *t*_10%_ when *D*(*t*) reaches 10% of the value that would have been reached at the same time if the exponential growth trajectory had continued. This is a measure of when the dynamics has significantly departed from the exponential regime. This point is located close to the peak, but is not biased by the slope of the exponential decline in the same way that the peak time is. The error of *t*_10%_ is determined again as the one standard deviation functional prediction interval.

We use the relative delay Δ*t* = *t*_10%_ − *t*_50%_ as a measure of the influence of public behaviour change on *β* and the rate of new exposure to the virus. The expectation for a PHI-driven scenario, versus an effect of a reduction in *S*, is that Δ*t* is comparable to the typical time from exposure to death, which is on the order of 20 days [20, 21].

### Corrected death counts

For COVID-19 dynamics, the attributed daily death rates are a more reliable metric than the number of confirmed cases, as they are not dependent on testing practices, and should be less prone to under-reporting. However, daily death rates are still subject to variations in reporting methods and in how each region defines what is classified as a COVID-related death [37]. Alternatively, *excess deaths* can be calculated by examining how many additional deaths (from all causes) have occurred above a baseline value that would be expected for the same time of year had the epidemic not happened [10]. While excess deaths statistics include deaths indirectly related to COVID19 (for example, ‘collateral’ deaths due to healthcare systems being overwhelmed, or reduced visits to emergency departments), and the baseline value for expected deaths may not exactly reflect the current situation [38] (for example, in lockdown conditions the number of road traffic accidents will be lower than typical historical values), the number of excess deaths is nevertheless widely agreed to be the most reliable indicator to reflect the state of the epidemic, and alleviates some of the shortcomings associated with the reported daily deaths data [37, 39].

We compare the time evolution of *D*(*t*) (aggregated to weekly level) to that of weekly excess deaths during the spring of 2020 relative to the median value of the historical data up to the past five years, where available [40]. Accounting for the delay that it takes in processing and registering the deaths in each case, we apply a constant correction factor to the reported daily deaths. These factors faithfully reflect the registered excess deaths, and by area were 1.9 (Italy), 2.2 (England), 1.4 (Spain), 1.5 (US), and we found that these same factors applied equally well to each of the corresponding metropolitan regions, respectively. For the purpose of fitting analysis, we continue to work with the scaled reported deaths because these data are available at a daily granularity, whereas the excess deaths are only provided weekly. The scaling of the data is shown in Fig. 3 (inset) for the example of England.

### SIR model with time-varying *β*(t)

Substantial efforts have gone into describing and predicting epidemic dynamics of many diseases, especially of COVID-19, through Monte Carlo-type discrete [41–43] or continuous differential equation models [44, 45]. Typically, these models require many input parameters such as fine-grained demographic and populations’ behavioural details. They are often critically dependent on these parameters with corresponding large variations between studies in terms of predicted outcomes and inevitable divergence from actual observations made after the forecasts [46]. In our work we instead aimed to understand the reduction of *S* and its impact in a largely model-independent way. In order to still derive quantitative trends and estimates, we chose the SIR model as the simplest epidemic model that takes time-dependent susceptibility (immunity) into account.

The well-established SIR framework is a deterministic model using a system of coupled ordinary differential equations, and is based on the compartmentalisation of populations into individuals who are susceptible, *S*, infectious, *I*, and no longer susceptible (removed), *R*. The *R* compartmentincludesindividualswhohave recovered from disease and survived, along with those who have died. The rate coefficients *β* and γ describe the flow from the *S* to the *I* and from the *I* to the *R* compartments, respectively, and the evolution of the system is described by the set of differential equations

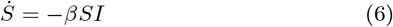

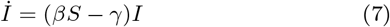

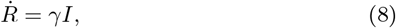

where the dot indicates a derivative with respect to time. These equations have been solved analytically for a constant *β* in [47].

Since public policy interventions and public behaviour changes (whether or not they are influenced by these interventions) affect the value of *β*, we introduce a time dependence of that parameter into the model *β* = *β*(*t*). Since no known intervention affects the duration of infectiousness, we treat γ as constant. Following the empirically found functional shape of the public response as representatively measured through use of public transit, we also model *β*(*t*) as a sigmoid with

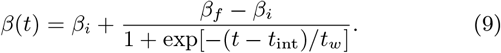

The shape of this function implies that there was only one period of relevant change to *β* from its unrestricted initial value *β_i_* to a later final value *β_f_*. This is justified as long as public policy relaxations do not take effect on *D*(*t*) and as long as compliance with them remains high (constant). We therefore restrict our main analysis to times that are not affected by such changes and only consider *D*(*t*) data until the end of May 2020, as no significant corresponding changes in TS(*t*) are discernible prior to early-May (considering the delay between exposure and death).

This simple model is based on several idealised assumptions. It does not account for inhomogeneity of the population or of the mixing dynamics [27], and so stochastic, regional and demographic effects are averaged out to *effective* parameters. In particular, it is known that *µ* is a parameter whose value strongly increases with age, but also with strong dependence on health status [48]. This means that not only the demographic composition of an investigated population needs to be considered, but also the dynamics of epidemic spread relative to it (e.g. the timing of seeding the virus in settings or population groups with specific demographic makeup such as schools, high-frequency business travellers or care homes). As a consequence, the infection fatality rates reported in the literature have varied, with some convergence in the range of 0.5% − 1% [49]. Findings outside the this range with potential country to country variations may still occur and be meaningful as effective average figures. Possible changes of *µ*(*t*) with time during the course of the epidemic have not been considered here, but are not likely to be important in the light of the high quality of exponential fits to the *D*(*t*) data. Since *S* is generally time-dependent over the course of the epidemic, we use a standard Runge-Kutta iterative algorithm to calculate numerical solutions of the system of differential equations [Eqs. (6) to (8)], and incorporate a time dependence to *β*(*t*) using Eq.9. A cost function is generated, derived from the mean-squared deviation between the numerical solutions for the epidemic trajectory and the observed data *D*_log_(*t*), in each geographical area considered. The individual cost functions for the metropolitan region and corresponding country are combined (with equal weighting) to simultaneously respect both sets of data in each pair, and a Nelder-Mead based optimisation method is then used to minimise this total cost function. This procedure provides our best-fit parameters for the epidemic dynamics.

Additionally in this process, coupling is introduced between each metropolitan region and its corresponding country by constraining the values of *β*(*t*) over time to be equal between the areas in each pair, as informed by the location data (Fig. 1b and Fig. 2 lower panels). Constraining the values of *β* in the metropolitan region and the rest of the country to be equal *β^m^*(*t*)= *β^c^*(*t*) is based on the assumption that public health interventions have equal effects on each area. This is supported by the observation that the time constants for initial exponential growth and the public behaviour changes (in timing and magnitude) are very similar for countries and their metropolitan regions following nationally homogeneously-imposed public health interventions, with the exception of the United States as already noted. The initial value of *β_i_* is determined by the previously determined (see above) exponential growth phase time constant. Free optimisation parameters then relate to timing of epidemic seeding and the nature of the change in *β* (*t*_int_, *t_w_*). To account for demographic differences, we use the distributions of ages from population statistics in each of the eight geographical areas along with the reported age-dependence of the IFR [20, 48] to calculate the average apparent IFR for each metropolitan region (*µ^m^*) and rest of country (*µ^c^*). Then, rather than constrain our model by exactly these determined IFR values (which may be prone to systematic bias in the clinical data), we instead couple the fitting procedure between metropolitan region and rest of country by prescribing the ratio *µ^m^*/*µ^c^* in each case. The obtained ratios used as constraints for each fit were 0.664 (London/England), 0.985 (Lombardy/Italy), 0.887 (Madrid/Spain), and 0.854 (New York/US), and so the metropolitan regions have a generally younger population structure (and therefore lower effective infection fatality rate). Finally, the absolute values of *µ* are then returned as fit parameters (see Extended Data), and are consistent with those reported in the literature [49]. An exception is the higher value obtained for Italy of *µ* =1.95%, which may be attributed in part to a comparably older population structure, and potentially the increased impact on healthcare capacities.

The model can be extended by incorporating further changes in *β*(*t*) as public policy interventions and/or compliance and public behaviour change. For the example of England and London we note that a slight increase of *β^c^* = *β^m^* =0.134 d^−1^ is consistent with the observations of *D*(*t*) from mid-May until the end of June 2020. The continued match of the data with equal values of *β*(*t*) while the exponential decay constants differ between London and the rest of England further strengthens the hypothesis that public policy interventions had an equal effect on *β* across both areas.

## Extended Data

**Table 2:** Exponential growth/decline time constants. Linear fits to the logarithm of daily deaths per million population data are used to avoid heteroskedastic bias. Fits to the initial growth phase for the total country level data in Fig. 1a produce time constant values which are all consistent with each other, with a mean of 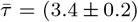 days. Country level data are then broken down into main metropolitan region and rest of country (Fig. 2), with corresponding fits to both the exponential growth and decline phases listed. Errors are 2 standard deviation confidence intervals (CI) from fits.

| Area | Initial Growth<br>$1/D_u$ (days) | Decline<br>$1/D_l$ (days) |
| --- | --- | --- |
| UK | $3.29 \pm 0.93$ | - |
| Rest of England | $4.22 \pm 0.83$ | $-22.0 \pm 1.0$ |
| London | $3.49 \pm 0.66$ | $-14.1 \pm 1.2$ |
| Spain | $3.0 \pm 1.1$ | - |
| Rest of Spain | $2.52 \pm 0.34$ | $-23.2 \pm 3.8$ |
| Madrid | $2.58 \pm 0.97$ | $-18.0 \pm 1.7$ |
| Italy | $3.43 \pm 0.77$ | - |
| Rest of Italy | $3.83 \pm 0.52$ | $-33.6 \pm 3.7$ |
| Lombardy | $3.5 \pm 1.7$ | $-24.3 \pm 3.7$ |
| US | $3.72 \pm 0.50$ | - |
| Rest of US | $4.07 \pm 0.95$ | $-56 \pm 27$ |
| New York | $2.72 \pm 0.37$ | $-15.7 \pm 0.9$ |

## Footnotes

1 ‘Recovering’ is defined here as ceasing to be infectious (and susceptible).

2 Unless otherwise indicated, all stated errors correspond to statistical one standard deviation uncertainties.

3 In Italy, early death reports were only available at the regional level; the population of Lombardy is dominated by Milan, which is the country’s largest metropolitan region.

4 For metropolitan versus rest-of-countrycomparisons, we usedmortality data for England with the assumption that public interventions for London best match those of England over the whole of the UK.

5 Threshold chosen in accordance with infection levels in countries with well-developed public health infrastructure (e.g. New Zealand, South Korea, Cuba, and Fiji) with proactive testing and ‘track and trace’ interventions resulting in lasting containment approaching elimination of locally-acquired cases.

